# Application of Evidence into Practice in Trauma: The Purpose to Practice (P2P) Approach to Consensus Generation

**DOI:** 10.1101/2025.08.20.25334072

**Authors:** Jenny M. Guido, Morgan Krause, Brendon Frankel, Emily Hillmer, Ariel W. Knight, Katheryn Grider, Ashley N. Moreno, Lacey N. LaGrone, Pamela Bixby, Stephanie Bonne, Eileen M. Bulger, James G Cain, Jennifer Chastek, Julia Roberts Coleman, Todd W Costantini, Nicholas Cozzi, Kimberly A. Davis, Rochelle A. Dicker, Warren C. Dorlac, Erik Van Eaton, Evert Eriksson, Susan Evans, Shannon Marie Foster, Jeffrey M. Goodloe, Elliott R. Haut, Molly Jarman, Alyssa Johnson, Meera Kotagal, Morgan Krause, John C. Kubasiak, Kelly Lang, Allison Barbara Leigh, Halinder S. Mangat, Debra Marie Marvel, Christopher Paul Michetti, Vicki Moran, Ashley N. Moreno, Simon JW Oczkowski, Michael A. Person, Michelle A. Price, LJ Punch, Megan Racey, Bradford L. Ray, Diane Redmond, Linda Kate Reinhart, Heather Rhodes, Bryn Rhodes, Andres M. Rubiano, Sabrina Sanchez, Babak Sarani, Erica Shelton, David A Spain, Kristan Staudenmayer, Deborah M. Stein, Julie Valenzuela, Cynthia Lizette Villarreal, Jeffrey L. Wells, Gabriela Zavala Wong, LeAnne Sitari Young

**Author notes:** Serving as Joint First Authors, Guido and Krause contributed equally to this paper. Corresponding Author: Lacey N. LaGrone, MD, MPH, MA 2500 Rocky Mtn Ave; Loveland, CO 80525 Trauma Acute Care Surgery, Medical Center of the Rockies, Loveland, CO, USA UCHealth; University of Colorado.

## Abstract

**Introduction:** In trauma care, there is a need to increase communication to ensure evidence-informed, best practice care guidelines are easily accessible to all providers to yield continuity of care. Clinical guidance use is one way to address this need while employing a patient-centered team approach.

**Methods:** During year two of the Design *for Implementation: The Future of Trauma Research & Clinical Guidance* conference series, participants gathered in person and virtually to further develop the Minimum Viable Product (MVP) created during year one. Professional facilitators used the purpose-to-practice (P2P) framework to help structure and guide further consensus building.

**Results:** Seventy in-person and up to 65 virtual attendees participated. Sixty-five responses were collected on the MVP reflection and initial feedback survey. Themes were developed surrounding the pillars of “Purpose,” “Principles,” “Participants,” and “Practices” while looking at the “Structure” for “Sustainability.” The “Purpose” pillar addressed the importance of rigorous, standardized implementation guidance. “Principles” exemplified the necessity of a collaborative approach and included all relevant stakeholders. Similarly, the central theme emphasized by the “Participants” pillar was the inclusiveness of all members of the trauma team. “Practices” dove into the deliverables of the initiative, including up-to-date decision-making support and logistics regarding guidance storage, management, and maintenance. Regarding “Structure,” the most highly ranked idea was developing a steering committee whose purpose would be primarily to prioritize strategic initiatives.

**Discussion:** Clinical guidance needs to be current and readily available to all providers. Next steps of this initiative include developing a steering committee and subcommittees to sustain momentum.

**What is already known on this topic –** The current state of trauma clinical guidance warrants optimization as clinicians experience implementation challenges, including relevancy to practice setting, inadequate access, and inadequate dissemination. Lack of guidance consensus and applicability highlights a need to equitably provide evidence-based recommendations to all providers to provide quality care for traumatically injured patients.

**What this study adds** – *This study models the use of a formal consensus-generation approach to optimize inclusivity of perspective integration and impact of output*.

**How this study might affect research, practice or policy** – *Recommended next steps for P2P consensus generation include the development of a steering committee and subcommittees to continue the work from this conference series to improve collaboration and accessibility of trauma clinical guidance nationally*.

**TSACO Manuscript Type:** Original Research

**LEVEL OF EVIDENCE:** VII

## INTRODUCTION

Comprehensive trauma care must be patient-centered and guided by best available evidence. Clinical guidance directly impacts care provision and outcomes, and thus must be current, concise, and patient-focused.^1^ Implementation and utilization of clinical guidance can support standardization and quality care provision, improving patient safety.^2^ Implementation of evidence-based recommendations is a long process with a mean of 17 years’ time to change clinical practice.^3^ Thus, few evidence-based interventions are ultimately incorporated into routine clinical practice.

Clinical guidance positively influences patient outcomes, but dissemination and implementation are often challenging and only variably successful.^4^ Individual providers may trust their experience over a guideline, may question the temporal relevance of a guideline, or feel that guidelines are not applicable to their practice setting due to resource constraints. All of these factors are barriers to successful implementation and highlight that guidelines often fall short by offering univariate solutions to a multivariate world.^5^ Current strategies to bridge this gap are fragmented, as various professional societies develop and distribute content independently rather than collaboratively. Additionally, for-profit vendors create niche widgets for clinical decision support tools. Thus, increased communication and collaboration between professional societies will improve accessibility of information to all providers and increase opportunities for continuity of holistic, patient-centered care.

One common solution to these barriers is formal consensus generation, in which a systematic process synthesizes key stakeholders’ and subject matter experts’ (SMEs’) input with best available evidence to generate a novel solution or recommendation. Commonly utilized across medical and scientific research, this process involves structured, formal engagement methods including Delphi techniques, consensus workshops, focus groups, and stakeholder advisory committees to ensure that the final product is not only robustly evidence-based but also accessible to and reproducible across differently resourced settings.^6,7^ Rooted in social science, psychology, decision theory, and systems thinking, formal engagement and consensus building aim to optimize transparency, inclusivity, and legitimacy in complex, dynamic decision-making processes.^7^ Effective formal engagement and consensus building have been shown to improve cooperation, legitimacy, and quality of proposed solutions to complex, dynamic issues.^8^

## METHODS

During the inaugural 2024 *Design for Implementation: The Future of Trauma Research & Clinical Guidance* (DFI) conference, conference attendees developed a Minimum Viable Product (MVP) to define the minimum level of success for this initiative.^1^ The deliverables attached to each component of the MVP are provided in **Table 1**. At the MVP’s core is the goal to relieve human suffering by improving the creation, dissemination, and implementation of trauma clinical guidance.

**Table 1.** Minimum Viable Product (MVP) Deliverables

| MVP Deliverable | Objective of the Deliverable |
| --- | --- |
| 1. Central repository for clinical guidance | The main objective of this MVP deliverable is to establish a central repository for clinical guidance. There needs to be electronic health record (EHR) interface capability with site-specific needs. |
| 2. Collaboration over Competition | A major goal of this MVP deliverable is to collaborate, not compete. There are many professional societies within trauma and surgery. Thus, it is essential to have the buy-in from all the communities involved with, and impacted by, guidance creation. Intentional efforts are made to ensure members of these societies are invited to participate in this conference series or to act as liaisons. Additionally, the need to foster a global health approach (versus an isolated approach) has been identified. |
| 3. Adaptive Clinical Guidance | This MVP deliverable focuses on adaptive clinical guidance and the need to stratify intervention recommendations based on the availability of resources.<sup>9</sup><br>The need to harmonize trauma patient management worldwide is essential but can be challenging due to the lack of consensus on protocols and guidelines. Clinical practice guidelines can become redundant or conflicting, and there can be different recommendations based on contrasting best practices. In turn, this becomes a bigger problem for low-resource trauma centers. This MVP deliverable requires a multidisciplinary approach to solve. |

### Purpose-to-Practice (P2P) Framework

Consensus building is critical to clinical guidance development as it facilitates deeper discussion among SMEs and other stakeholders.^7^ This was facilitated via the purpose-to-practice (P2P) framework. This is a structured approach to ensure that organizations and key stakeholders’ core values and objectives directly inform decision-making processes and eventual practices.^11^ P2P was selected for the particular initiative given its applicability to early-stage projects that require structural development, its ability to engage a larger number of stakeholders who are allowed equal contributions, and its promotion of consensus building via group conversation and idea generation.^11^ Furthermore, it is a cost-friendly approach that is feasible for stakeholders to attend in a series of shorter sessions over weeks to months. In this format, stakeholders design their new initiative around five predefined elements of success: *Purpose*, *Principles*, *Participants*, *Practices*, and *Structure*, as presented in **Figure 1**. In regards to “Purpose,” participants consider the importance of their work to themselves and their communities. Rules of engagement and measures of success are defined in “Principles.” “Participants” are clearly defined stakeholders inherent to achieving the former two pillars, who are then organized at macro-and micro-levels to achieve the common purpose via “Structure.” Finally, “Practices” focuses on dissemination and implementation. Ultimately, via early collaboration, a successful product aligns with stakeholders’ priorities and appropriately serves their partner groups by maintaining focus on an initiative’s critical areas. In this initiative, for example, the MVP served as the “Practice” prototype for redesign of national trauma clinical guidance.

**Figure 1.**
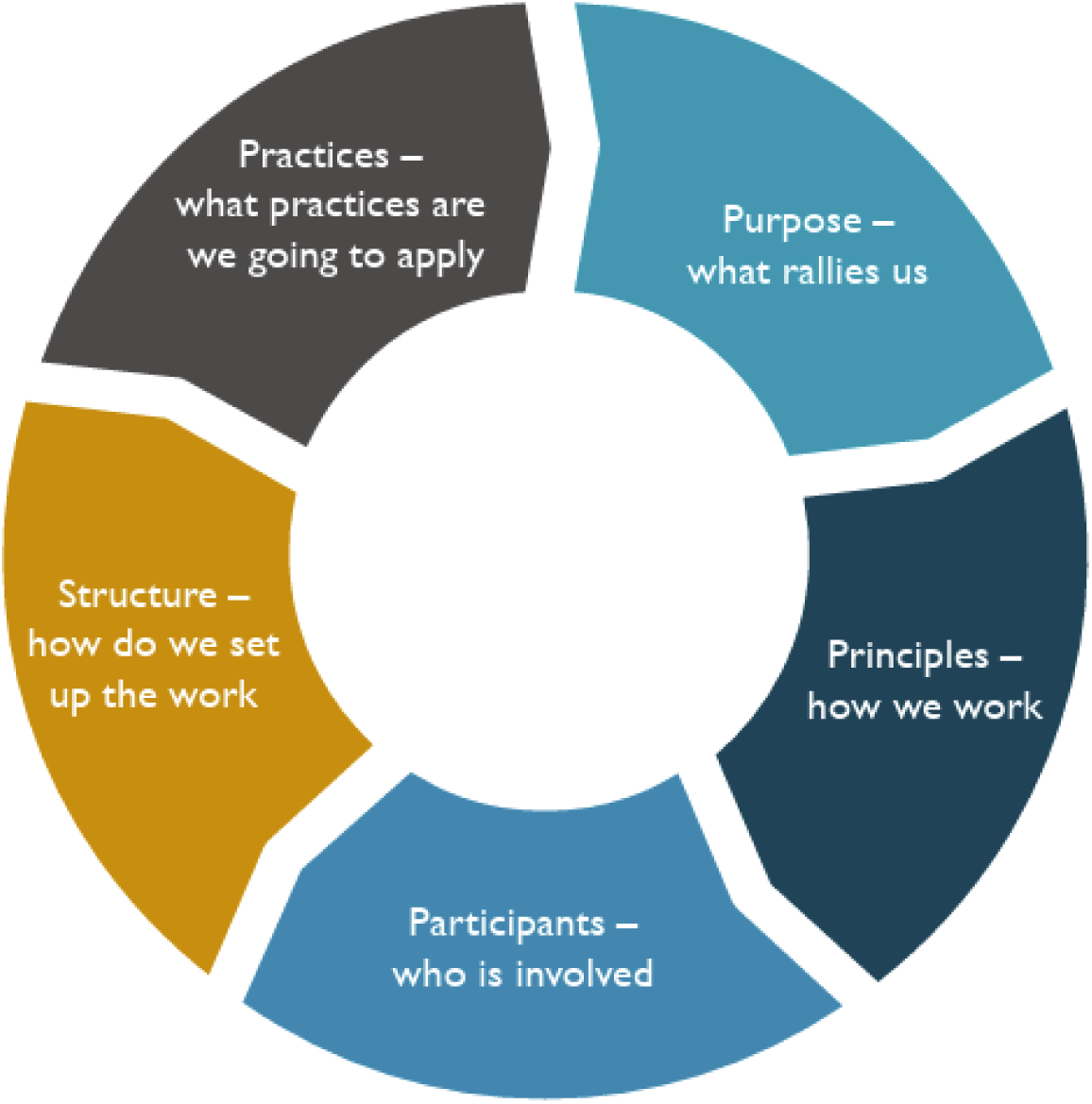
The Five Elements of the Purpose-to-Practice Framework

### MVP Reflection and Initial Feedback

Following presentation of the P2P framework, participants completed an initial qualitative Microsoft Forms (Microsoft 365, Version 2025; Redmond, WA) survey regarding its “Purpose,” “Principles,” “Participants,” and “Practices” elements. Details of the survey prompts as they relate to the MVP are presented in **Table 2**. The essence of this MVP reflection centered on practices to be applied, their benefits to end-users, and implementation strategies.

**Table 2.** MVP Reflection and Initial Feedback Using the Purpose-to-Practice (P2P) Framework

| Thematic Prompt Under Review | Prompt-to-Prompt (P2P) |
| --- | --- |
| **Purpose:** Relieve human suffering by improving trauma clinical guidelines design and implementation. In the United States, an injured person seen at the worst performing trauma center is twice as likely to die as a similar patient seen at the best performing trauma center. The National Academies of Sciences Engineering and Medicine Zero Preventable Deaths report identified application of evidence-based medicine within learning health systems to be a key opportunity for improvement to close these outcome gaps. The vast majority of providers caring for injured patients do not have access to relevant, timely clinical information that would allow them to practice evidence-based medicine, thus exacerbating outcome inequity and contributing to preventable deaths. | Why is the work important to you and the larger community?<br>As you consider the prompt above, do you see anything missing from this statement/MVP output? |
| **Principles:** Collaborative – Working together, generously and transparently, on shared goals and outputs. Inclusive – All trauma care providers and patients are part of the design and implementation process, including diverse practice types, community identification, and geographic locations. Accessible and Useful – Free, timely, current, and formatted for easy bedside use. | What rules must we absolutely obey to succeed in achieving our purpose? (aka: Must do, must not do)<br>As you consider the prompt above, do you see anything missing from this statement/MVP output? |
| **Participants:** Primary users and therefore participants: - Whoever is providing trauma care to the patient: (EM, APP, Gen. Surg, Low Volume Trauma, Nurses, Pre-Hospital)- Trauma Surgeons - High Volume - Pediatrics - Patients<br>Secondary users and therefore participants:<br>- Associations - Hospital Leader/Administration - Policymakers/Regulators - Payers / Funders (NIH, AHRQ, State, Prof. societies, institutions)<br>Diversity of participants is critical to success, and we will actively include participants of diverse practice types, community identification and geographic location. | Who can contribute to achieving our purpose and must be included?<br>As you consider the prompt above, do you see anything missing from this statement/MVP output? |
| **Practices:** MVP Objectives were prioritized as follows: 1) Develop an accessible (free/offline), easy-to-use platform to house existing guidelines for use in every hospital, every time, every patient. 2) Ensure guidelines on the platform are consolidated, coordinated (e.g., not competing) and current. 3) Build a consensus around a standard for future guidelines, and a use-case test with 1 – 2 clinical scenarios that follow criteria established by the group. 4) Evolve prioritized guidelines to deliver stratified guidance by | What are we going to do? What will we offer to our users and how will we do it?<br>As you consider the prompt above, do you see |
| resource environments. 5) WILD CARD: Catalyze better data collection and integration (social and clinical). | anything missing from this statement/MVP output? |

Univariate statistical and thematic analyses of grouped survey responses were then conducted using Microsoft Excel (Microsoft Corporation, 2018; Redmond, WA). Themes were deemed significant when they appeared in at least two responses within a pillar.

### Scalability and Sustainability Session

From here, individual and small group sessions were held to focus on the “Structure” element of the P2P framework. In this process, participants reflected individually, in pairs, and in small groups to address the question, “How must we organize to distribute control and achieve our purpose?” While both in-person and virtual participants were encouraged to reflect individually, only in-person participants joined together in pairs and small groups. Each small group then shared back their ideas on how to creatively organize stakeholders while facilitators recorded any common themes that emerged. Participants then used a dot voting system to vote for up to three themes they deemed most important. Common themes of successful stakeholder organization were collected and subsequently ranked. A thematic analysis was then performed with Microsoft Excel (Microsoft Corporation, 2018; Redmond, WA). Predominant themes, detailed in the Results section, were deemed significant when they appeared in at least two responses within a pillar.

### Post-Conference Survey

Finally, a standardized, anonymous post-conference survey was conducted using Microsoft Forms (Microsoft 365, Version 2505; Redmond, WA). Respondents were additionally asked the open-ended question, “What do you see as the main barrier to ongoing collaboration/productivity of the DFI initiative?” Univariate descriptive statistics and thematic analysis were conducted.

The Consensus-Based Checklist for Reporting of Survey Studies (CROSS) was used to help report the two surveys (Supplemental Item 1).^12^

## RESULTS

### Demographics of Conference Participants

The second annual DFI conference brought together 70 in-person and up to 65 virtual participants. Demographics were captured in the post-conference survey (n=56). Most attendees were clinicians (46% physicians, 16% nurses, 4% advanced practice providers), most of whom (75%) worked in either trauma or general surgery or emergency medicine. A complete breakdown of the respondent demographics is available in Supplemental Item 2.

### MVP Reflection and Initial Feedback

A total of 62 attendees provided survey feedback on the MVP. Of these respondents, “n” denotes the number of participants who indicated a missing element from the corresponding P2P statement. Pillar-specific themes are further discussed below.

#### Purpose (n=16, 25.8%)

Multiple barriers to the “Purpose” statement were identified. Concerns were raised about the feasibility of guidance implementation due to local resource constraints. As one respondent noted, implementation leaders must account for the *“limitations of some hospitals regarding what is needed to carry out*” specific guidelines. Thus, additional stakeholders, including rural and community hospitals in addition to national and global professional societies and SMEs, warrant inclusion for successful guidance design and implementation to ensure realistic applicability between differently resourced settings. Trauma center level designation and scope of practice, urban versus rural location, and individual community factors must be considered in creation of universally applicable clinical guidance. Many respondents highlighted nonadherence to current clinical guidelines and lack of a mechanism to enforce standards of care as barriers to “Purpose.” Finally, a few respondents noted that guideline adherence may depend upon the time of care delivery relative to the time of injury (i.e., prehospital trauma care requires different guidance and implementation strategies compared to postoperative trauma intensive care provision).

#### Principles (n=14, 22.5%)

All respondents emphasized the importance of stakeholder inclusion in “Principles” with particular mention of clinicians as well as hospital and community support staff, rehabilitation specialists, and medical logistics planners. Respondents also highlighted that clinical guidance development should take into account the applicability to both higher and lower resourced environments. Identified areas of concern included financial sustainability, inclusion of payors as stakeholders, and anticipation of required guideline update and maintenance. Lastly, one respondent proposed a strategy of engaging only with key constituents upfront and expanding opportunities for additional stakeholder input in later phases of guideline development given hypothetical difficulties of upfront inclusion of all relevant stakeholders.

#### Participants (n=18, 29.0%)

Respondents identified several improvements to the “Participants” pillar, most notably ensuring inclusion of the entire spectrum of clinicians, medical staff, and caregivers in the guidance development process. Participants cited the importance of engaging allied health professionals and community-based service providers such as physical therapy, respiratory therapy, medical logistics planners, researchers, quality improvement personnel, and caretakers as well as other surgical subspecialists and non-trauma clinicians who also provide care to injured patients. Further engagement of professional societal leadership was identified as an opportunity to further such multidisciplinary collaboration.

#### Practices (n=16, 25.8%)

With respect to “Practices,” most respondents highlighted the technical challenges of clinical guidance development, including decision aids as well as guidance maintenance, updates, storage, and management logistics. Respondents emphasized a need to better standardize and integrate guidance between all professional trauma organizations. Care variability between differently resourced settings was again highlighted as a variable, with particular focus on feasible alternative strategies to be considered when the best treatment options are not available.

### Scalability and Sustainability Session

Respondents emphasized the development of a steering committee to prioritize strategic initiatives as a critical step to maintain this clinical guidance initiative (Figure 2). This was closely followed by the establishment of subcommittees to focus on various topics including guideline development, scientific research, and implementation and dissemination strategies. Respondents also recommended that professional trauma societies consider adoption of the National Comprehensive Cancer Network’s (NCCN) model in which multiple guidelines are simultaneously developed, maintained, and updated.^13^ Lastly, additional responses highlighted the need to identify the right collaborative partners and maintain certain data standards.

**Figure 2.**
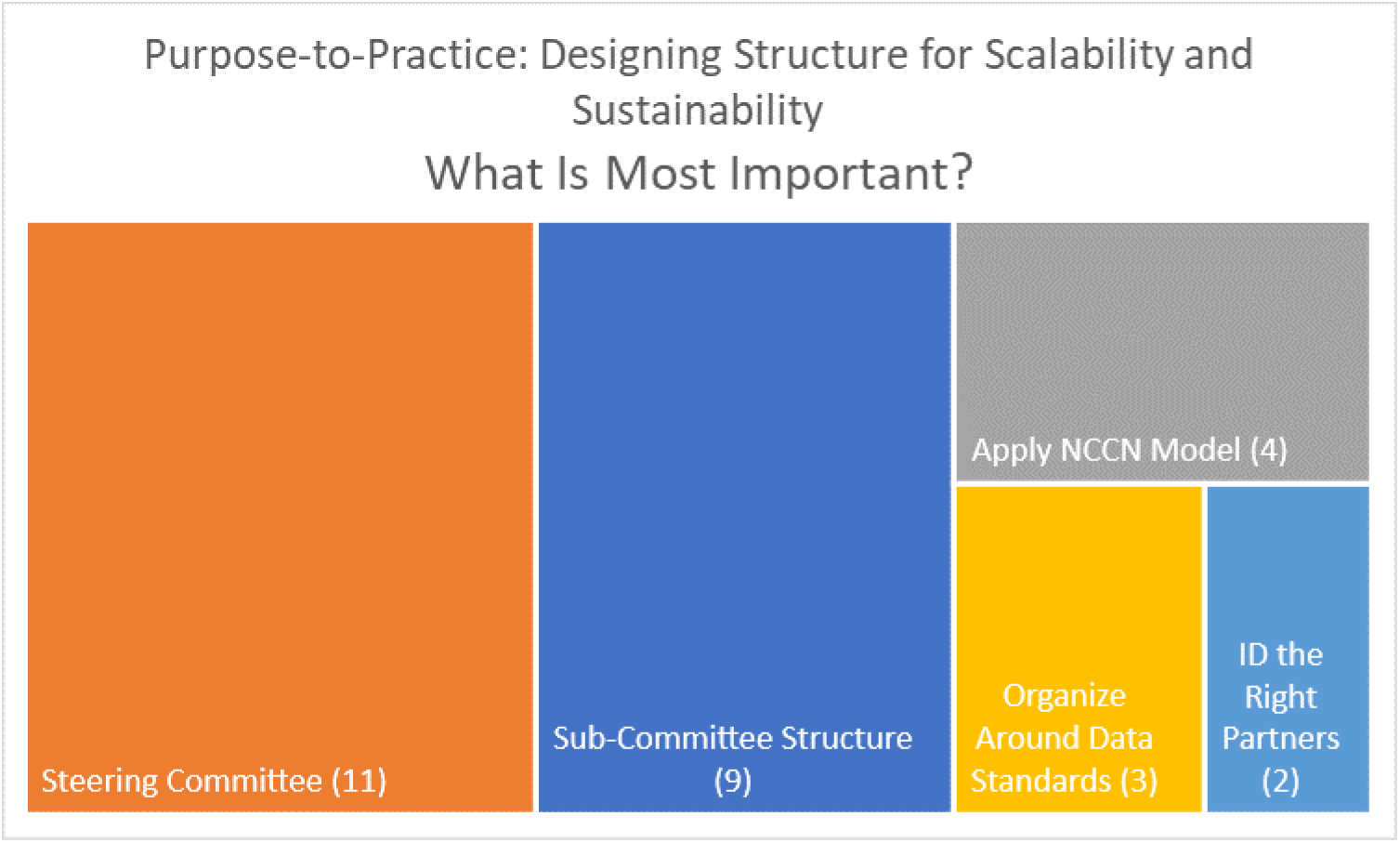
Results of the dot voting exercise and areas of most importance

### Barriers to Ongoing Collaboration

Several themes were identified as barriers to ongoing collaboration and productivity, most notably lack of reliable funding to not only build, implement, and disseminate comprehensive guidance, but also to sustain its maintenance and development in a way that is applicable to the broadest possible audience of trauma care providers. Respondents also identified engagement, or lack thereof, as another such barrier, noting that without regular participatory intervals, only minimal progress will be made on the MVP and respective deliverables. Formation of dedicated subcommittees that regularly meet throughout the year allows for ongoing interpersonal collaboration and more sustainable clinical guidance development and refinement through identification and achievement of smaller goals over time. It is imperative, however, to balance cohesiveness, subcommittee tasks, and time commitments with individuals’ other responsibilities to ensure members maintain a level of engagement with their peers and focus on the MVP. In addition, differing long-term goals may be a barrier to collaboration. As many surgeons rely on academic productivity and incentives for professional advancement, some of these benefits may be lost via support of common trauma clinical guidance development given relatively fewer unique opportunities to publish. The willingness or lack thereof of professional societies to collaborate may also act as a barrier. Lastly, adequate representation from all national trauma organizations and quality improvement programs is imperative to creation of a centralized authority and engendering widespread trust amongst their respective communities.

## DISCUSSION

The DFI conference series was founded to engender collaboration between professional trauma societies to create a shared central repository for standardized, evidence-based, contemporary clinical guidance in the care of injured patients. The P2P framework was instrumental in further refining the MVP for this initiative. Its five elements, “Purpose,” “Principles,” “Participants,” “Practices,” and “Structure” highlight several key themes imperative to the initiative’s success and sustainability. The importance of successfully developing rigorous, evidence-based, best practice trauma guidelines hinges on several concepts: engagement of key stakeholders throughout development, inclusion of all providers involved in the spectrum of trauma care, consideration of the universal applicability of guidance based upon variations in locally available resources, measures to assess guideline adherence and standardization, and careful consideration of successful guideline accessibility, dissemination, and implementation. Ultimately, SMEs supported the development of a steering committee to oversee several diverse subcommittees dedicated to each of the aforementioned topics. Lastly, it is imperative to have buy-in from all major trauma societies to engender an appropriate level of professional trust in a centralized repository of clinical guidelines. Of note, many of the technical aspects of clinical guidance and development have been addressed in other DFI sessions (*please refer to Zavala-Wong et al. 2025 and Frankel et al. 2025 of this supplement – manuscripts submitted; under consideration*).^14,15^

The P2P framework discussion informed the development and prioritization of three MVP deliverables.

### MVP Deliverable #1 – Central Repository for Clinical Guidance

Creation of bedside-ready clinical guidance materials that are immediately available and readily accessible is imperative to improve standardization of trauma care and its delivery across widely varying healthcare settings. When surveyed during the DFI conference, 89% of respondents indicated support for a mobile application to serve as a repository of evidence-based, best available practices to supplement a local provider’s clinical judgment. The other 11% of respondents expressed concern over the readiness of such a mobile application (please refer to Frankel et al. 2025 of this supplement – manuscript submitted; under consideration).^15^

Nevertheless, successful implementation of mobile applications as supplements to clinical judgment and modes of continuing education has been established in other medical specialties. The overwhelming majority of healthcare professionals use a smartphone and up to half utilize medical applications in clinical care.^16,17^ Given such widespread use, development of a mobile application to act as a clinical guidance repository is likely to be readily accessible to many trauma care providers. Notably, mobile applications touting clinical information have already been developed for use in other specialties, including oncology and dermatology.^18–20^ While such resources indeed improve clinicians’ and patients’ access to information and encourage further knowledge acquisition, several limitations exist.^20^ Notably, cost of either an application or additional content within an application is often prohibitive.^20^ Furthermore, development of such an application must be done thoughtfully and deliberately as a lack of evidence-based recommendations not only undermines the quality of care being delivered but also trust in and reputation of professional societies.^16,17,20^ These findings reinforce the respondents’ feedback that any clinical guidance repository be available, accessible, evidence-based, and endorsed by professional societies to ensure equitable dissemination and informational quality control.

### MVP Deliverable #2 – Collaboration over Competition

Collaboration between individuals as well as professional societies is imperative to successful clinical guidance creation and dissemination. At present, individual professional trauma societies publish separate, often discrepant guidelines on similar topics, thus limiting standardization of best clinical practices in the more widespread care of injured patients. The collaborative leadership of organizational stakeholders and trauma SMEs is essential to these efforts’ success.

Development of a steering committee to lead such efforts was identified as a sustainability measure. Notably, the Coalition for National Trauma Research’s (CNTR) Evidence-to-Practice (E2P) Committee formed as a result of the DFI conference. The E2P Committee has representatives from major professional trauma societies as well as several diverse at-large members, and will serve as an advisory group to oversee the research, development, dissemination, and implementation of clinical guidance efforts. Similar collaborations between other professional medical organizations and SMEs from different clinical backgrounds, including medical/surgical specialty and country of practice, have helped standardize recommended best practices for commonly encountered clinical problems, most notably atrial fibrillation, but also pulmonary embolism and COPD.^21–24^ However, the dissemination and implementation of these recommendations remains less clear. To date and to the authors’ knowledge, no such collaboration has occurred on as large a scale as we are proposing nor for such a vast range of topics, but these previous collaborations demonstrate the feasibility of the development of truly comprehensive, evidence-based guidelines.

### MVP Deliverable #3

Adaptive Clinical Guidance – Nation- or worldwide standardization of trauma care is a particular challenge to consensus guideline development, especially for lower resourced centers and environments. Furthermore, as previously discussed, it is essential to resolve discrepancies between different societal recommendations to avoid confusion and to minimize variability. Multiple barriers that limit working relationships between professional trauma societies were identified along with potential solutions. However, seven major national family medicine societies were able to successfully collaborate to clearly define a common new practice model, improve access to equitable care, optimize practice finances, implement new technologies, and provide evidence-based continuing education for their specialty.^25^ While the authors’ scope did not specifically include clinical guidance, their work clearly demonstrates the ability of national professional organizations to productively work together.

This work has several limitations. Firstly, the key partners and SMEs who participated in the DFI initiative all have personal and/or professional interests in these topics and the associated subsequent work, potentially creating unconscious biases. Furthermore, the perspectives and suggestions shared throughout this consensus-generating process may neither reflect the needs of or be applicable to trauma clinicians in different practice environments with widely variable resources both domestically and internationally.

## Conclusion

Use of the P2P framework to support engagement of key stakeholders and SMEs to optimize access to and dissemination of a central repository of clinical trauma guidance was successful. Overall, these comments highlight that formal engagement methods provide a predefined structure for stakeholders to collaborate and achieve consensus on the management of complex problems. Further refinement and ongoing assessments will be key to the development of a sustainable workflow and ongoing successful interorganizational collaboration.

## SUPPLEMENTAL ITEMS

Supplemental Item 1. Consensus-Based Checklist for Reporting of Survey Studies (CROSS)

Supplemental Item 2. Demographics

## AUTHOR CONTRIBUTION STATEMENT

Design: KG, ANM, LNL, and the 2025 Design for Implementation (DFI) Authorship Group

Data acquisition: BF, KG, ANM, LNL, and the 2025 Design for Implementation (DFI) Authorship Group

Analysis and interpretation: JMG, MK, BF, EH, KG, ANM, LNL

Drafting and Critical Revision: JMG, MK, BF, EH, KG, ANM, LNL, AWK, and the 2025 Design for Implementation (DFI) Authorship Group

Guarantor: LNL

## FUNDING STATEMENT

The Design for Implementation: The Future of Trauma Research & Clinical Guidance (DFI) conference series was made possible, in part, by a conference grant from the Agency for Healthcare Research and Quality (1R13HS028940-01A1). The views expressed in written conference materials or publications and by speakers and moderators do not necessarily reflect the official policies of the Department of Health and Human Services; nor does mention of trade names, commercial practices, or organizations imply endorsement by the U.S. Government.

The American Association for the Surgery of Trauma, American Burn Association, American Trauma Society, Eastern Association for the Surgery of Trauma, Chest Wall Injury Society, Society of Critical Care Medicine, Society of Trauma Nurses, and Trauma Center Association of America provided travel funds for representatives to attend the 2025 DFI meeting. The Eastern Association for the Surgery of Trauma and American Trauma Society provided travel funds for patient partners to attend. The Society of Trauma Nurses, Acera Surgical, and Tactuum provided additional financial support. The American College of Surgeons hosted the event in its Chicago offices and provided meeting management, facilities, and audiovisual equipment at no cost to the conference.

Publication of this supplement is made possible by Medical Center of the Rockies, UCHealth (Loveland, Colorado, United States).

## HUMAN SUBJECTS STATEMENT

This study was reviewed by the Colorado Multiple Institutional Review Board, CB F490; COMIRB No: 24-1608 and COMIRB #: 22-0626, and determined exempt from institutional review board review.

## Data Availability

A limited dataset of deidentified data produced in the present study are available upon reasonable request to the authors.

---

Checklist for Reporting of Survey Studies (CROSS)

Checklist for Reporting of Survey Studies (CROSS)
| Section/topic | Item | Item description | Reported on page # |
| --- | --- | --- | --- |
| **Title and abstract** |  |  |  |
| Title and abstract | 1a | State the word “survey” along with a commonly used term in title or abstract to introduce the study’s design. | n/a |
|  | 1b | Provide an informative summary in the abstract, covering background, objectives, methods, findings/results, interpretation/discussion, and conclusions. | Abstract |
| **Introduction** |  |  |  |
| Background | 2 | Provide a background about the rationale of study, what has been previously done, and why this survey is needed. | 13 |
| Purpose/aim | 3 | Identify specific purposes, aims, goals, or objectives of the study. | 13 |
| **Methods** |  |  |  |
| Study design | 4 | Specify the study design in the methods section with a commonly used term (e.g., cross-sectional or longitudinal). | 14, 17 |
|  | 5a | Describe the questionnaire (e.g., number of sections, number of questions, number and names of instruments used). | n/a |
| Data collection methods | 5b | Describe all questionnaire instruments that were used in the survey to measure particular concepts. Report target population, reported validity and reliability information, scoring/classification procedure, and reference links (if any). | n/a |
|  | 5c | Provide information on pretesting of the questionnaire, if performed (in the article or in an online supplement). Report the method of pretesting, number of times questionnaire was pre-tested, number and demographics of participants used for pretesting, and the level of similarity of demographics between pre-testing participants and sample population. | n/a |
|  | 5d | Questionnaire if possible, should be fully provided (in the article, or as appendices or as an online supplement). | n/a |
| Sample characteristics | 6a | Describe the study population (i.e., background, locations, eligibility criteria for participant inclusion in survey, exclusion criteria). | n/a – conference attendees |
|  | 6b | Describe the sampling techniques used (e.g., single stage or multistage sampling, simple random sampling, stratified sampling, cluster sampling, convenience sampling). Specify the locations of sample participants whenever clustered sampling was applied. | n/a |
|  | 6c | Provide information on sample size, along with details of sample size calculation. | 17 |
|  | 6d | Describe how representative the sample is of the study population (or target population if possible), particularly for population-based surveys. | n/a |
| Survey administration | 7a | Provide information on modes of questionnaire administration, including the type and number of contacts, the location where the survey was conducted (e.g., outpatient room or by use of online tools, such as SurveyMonkey). | 15, 17 |
|  | 7b | Provide information of survey's time frame, such as periods of recruitment, exposure, and follow-up days. |  |
|  | 7c | Provide information on the entry process:<br>→For non-web-based surveys, provide approaches to minimize human error in data entry.<br>→For web-based surveys, provide approaches to prevent "multiple participation" of participants. | n/a |
| Study preparation | 8 | Describe any preparation process before conducting the survey (e.g., interviewers' training process, advertising the survey). | n/a |
| Ethical considerations | 9a | Provide information on ethical approval for the survey if obtained, including informed consent, institutional review board [IRB] approval, Helsinki declaration, and good clinical practice [GCP] declaration (as appropriate). | Title page |
|  | 9b | Provide information about survey anonymity and confidentiality and describe what mechanisms were used to protect unauthorized access. | 17 |
| Statistical analysis | 10a | Describe statistical methods and analytical approach. Report the statistical software that was used for data analysis. | 14, 17 |
|  | 10b | Report any modification of variables used in the analysis, along with reference (if available). | n/a |
|  | 10c | Report details about how missing data was handled. Include rate of missing items, missing data mechanism (i.e., missing completely at random [MCAR], missing at random [MAR] or missing not at random [MNAR]) and methods used to deal with missing data (e.g., multiple imputation). | n/a |
|  | 10d | State how non-response error was addressed. | n/a |
|  | 10e | For longitudinal surveys, state how loss to follow-up was addressed. | n/a |
|  | 10f | Indicate whether any methods such as weighting of items or propensity scores have been used to adjust for non-representativeness of the sample. | n/a |
|  | 10g | Describe any sensitivity analysis conducted. | n/a |
| **Results** |  |  |  |
| Respondent characteristics | 11a | Report numbers of individuals at each stage of the study. Consider using a flow diagram, if possible. | 17 |
|  | 11b | Provide reasons for non-participation at each stage, if possible. | n/a |
|  | 11c | Report response rate, present the definition of response rate or the formula used to calculate response rate. | n/a |
|  | 11d | Provide information to define how unique visitors are determined. Report number of unique visitors along with relevant proportions (e.g., view proportion, participation proportion, completion proportion). | n/a |
| Descriptive results | 12 | Provide characteristics of study participants, as well as information on potential confounders and assessed outcomes. | 17 |
| Main findings | 13a | Give unadjusted estimates and, if applicable, confounder-adjusted estimates along with 95% confidence intervals and p-values. | n/a |
|  | 13b | For multivariable analysis, provide information on the model building process, model fit statistics, and model assumptions (as appropriate). | n/a |
|  | 13c | Provide details about any sensitivity analysis performed. If there are considerable amount of missing data, report sensitivity analyses comparing the results of complete cases with that of the imputed dataset (if possible). | n/a |
| **Discussion** |  |  |  |
| Limitations | 14 | Discuss the limitations of the study, considering sources of potential biases and imprecisions, such as non-representativeness of sample, study design, important uncontrolled confounders. | 21 |
| Interpretations | 15 | Give a cautious overall interpretation of results, based on potential biases and imprecisions and suggest areas for future research. | 21-22 |
| Generalizability | 16 | Discuss the external validity of the results. | n/a |
| **Other sections** |  |  |  |
| Role of funding source | 17 | State whether any funding organization has had any roles in the survey's design, implementation, and analysis. | Conflicts of Interest section |
| Conflict of interest | 18 | Declare any potential conflict of interest. | Conflicts of Interest section |
| Acknowledgements | 19 | Provide names of organizations/persons that are acknowledged along with their contribution to the research. | n/a |
| Sharma A, Minh Duc NT, Luu Lam Thang T, Nam NH, Ng SJ, Abbas KS, Huy NT, Marušić A, Paul CL, Kwok J, et al. A Consensus-Based Checklist for Reporting of Survey Studies (CROSS). *J Gen Intern Med*. 2021;36(10):3179-3187. DOI: 10.1007/s11606-021-06737-1. |  |  |  |

**Supplemental Item 1.** Demographics of Post-Conference Survey Respondents (N=56)*

| Variable | N | PERCENT | Variable | N | PERCENT |
| --- | --- | --- | --- | --- | --- |
| Provider Type |  |  | Global health initiatives |  |  |
| Physician | 26 | 46.4% | Yes | 14 | 25.0% |
| Non-Clinical/Not Applicable | 10 | 17.9% | No | 38 | 67.9% |
| RN | 9 | 16.1% | Not Applicable | 4 | 7.1% |
| Other | 9 | 16.1% | Verified/designated trauma center |  |  |
| Advanced Practice Providers (APP) | 2 | 3.6% | Yes | 34 | 60.7% |
| Appointment |  |  | No | 9 | 16.1% |
| Academic | 29 | 51.8% | Not applicable | 13 | 23.2% |
| Non-academic | 19 | 33.9% | Trauma Center Level |  |  |
| Other | 8 | 14.3% | Level I | 24 | 70.6% |
| Gender |  |  | Level II | 5 | 14.7% |
| Woman | 31 | 55.4% | Level III | 3 | 8.8% |
| Man | 23 | 41.1% | Level IV | 2 | 5.9% |
| Transgender | 1 | 1.8% | Clinical discipline |  |  |
| Prefer not to answer | 1 | 1.8% | Trauma Surgery | 21 | 37.5% |
| Age |  |  | Emergency Medicine | 8 | 14.3% |
| 25-34 | 4 | 7.1% | General Surgery | 13 | 23.2% |
| 35-44 | 17 | 30.4% | Pediatrics | 1 | 1.8% |
| 45-54 | 18 | 32.1% | Burns | 1 | 1.8% |
| 55-64 | 17 | 30.4% | Orthopaedics | 1 | 1.8% |
| Race/Ethnicity |  |  | Neurology | 1 | 1.8% |
| White | 44 | 78.6% | Anesthesia | 3 | 5.4% |
| American Indian or Alaska Native | 1 | 1.8% | Physical and Rehabilitation Medicine | 1 | 1.8% |
| Asian | 5 | 8.9% | Neurosurgery | 2 | 3.6% |
| Black or African American | 4 | 7.1% | Internal Medicine | 1 | 1.8% |
| Hispanic | 1 | 1.8% | Other | 5 | 8.9% |
| Prefer not to answer | 1 | 1.8% | Not Applicable | 14 | 25.0% |
| Degree |  |  | Participating in dissemination science/quality improvement |  |  |
| Bachelor of Science and/or Arts | 8 | 14.3% | Yes | 28 | 50.0% |
| Bachelor of Science in Nursing | 3 | 5.4% | No | 23 | 41.1% |
| Master of Science and/or Arts | 8 | 14.3% | Not Applicable | 5 | 8.9% |
| Master of Science in Nursing | 7 | 12.5% | Conducts Research |  |  |
| Master of Business Administration | 6 | 10.7% | Yes | 29 | 51.8% |
| Master of Public Health | 7 | 12.5% | No | 25 | 44.6% |
| Doctor of Medicine | 26 | 46.4% | Not Applicable | 2 | 3.6% |
| Doctor of Philosophy | 7 | 12.5% | Participates in clinical guidance development |  |  |
| Other | 7 | 12.5% | Yes | 39 | 69.6% |
| None | 1 | 1.8% | No | 13 | 23.2% |
| Rural |  |  | Not Applicable | 4 | 7.1% |
| Yes | 7 | 12.5% | Context of clinical guidance development |  |  |
| No | 36 | 64.3% | International | 10 | 25.6% |
| Not Applicable | 13 | 23.2% | National | 23 | 59.0% |
| Military |  |  | Regional | 3 | 7.7% |
| Yes | 2 | 3.6% | Institutional | 22 | 56.4% |
| No | 46 | 82.1% | Other | 3 | 7.7% |
| Not Applicable | 8 | 14.3% | Role in Publishing |  |  |
| Patient Advocate |  |  | No/None | 18 | 32.1% |
| Yes | 13 | 23.2% | Associate Editor/Editor/Editorial Board | 13 | 23.2% |
| No | 38 | 67.9% | Regular Reviewer | 13 | 23.2% |
| Not Applicable | 5 | 8.9% | Other | 4 | 7.1% |
| Professional role in diversity, equity,<br>and inclusion (DEI) initiatives |  |  | Not Applicable | 8 | 14.3% |
| Yes | 9 | 16.1% |  |  |  |
| No | 41 | 73.2% |  |  |  |
| Not Applicable | 6 | 10.7% |  |  |  |

**\*Percentages over 100% are due to rounding error**

